# COVID-19 Vaccine Acceptance and Uptake Among Healthcare Workers in Trinidad & Tobago

**DOI:** 10.1101/2022.05.09.22274854

**Authors:** Chavin D. Gopaul, Dale Ventour, Davlin Thomas

**Author notes:** Corresponding Author: Dr. Chavin D. Gopaul, Ground Floor, Building 7, Eric Williams Medical Science Complex, North Central Regional Health Authority, Champs Fleur, Uriah Butler Highway, Trinidad. (868)645-2640 Ext. 2601.

## Abstract

**Background:** COVID-19 vaccine acceptance is important in ensuring the widespread vaccination of the population to achieve herd immunity. Establishing the acceptance of vaccines among healthcare workers, who play a vital role immunization program success, is important. The aim of this study was to assess the influence of social trust and demographic factors on COVID-19 vaccine acceptance among healthcare workers.

**Methods:** A cross-sectional survey utilizing an electronic questionnaire inquiring about COVID-19 vaccine uptake, preferences, and concerns was distributed via email to 1,351 North Central Regional Health Authority (NCRHA) healthcare workers of the following categories: medical practitioners, nursing personnel, veterinary surgeons, medical interns, dental interns, paramedics, and pharmacists. These professions were selected as they were granted power to administer COVID-19 vaccines during this period of public emergency by the President of Trinidad and Tobago and were therefore likely to be NCRHA healthcare workers directly involved in vaccine administration services. 584 participants returned a completed questionnaire. Bivariate analysis using Chi-square analysis of association was used to determine the association between the respondents’ characteristics and the acceptance of the vaccine and the association between vaccine acceptance among healthcare workers and trust. The association between the acceptance of the COVID-19 vaccines and healthcare workers’ characteristics and trust was established using multinomial logistic regression.

**Results:** A total of 584 healthcare workers took part in the study and 1.4% showed unwillingness to receive COVID-19 vaccine. The study indicates that age, profession, and the trust in international organizations and other healthcare providers predict the uptake of COVID-19 vaccines among healthcare workers. However, gender of the healthcare workers does not predict vaccine acceptance.

**Conclusions and Relevance:** Efforts towards enhanced vaccine acceptance among healthcare workers should take into consideration age, profession, and the trust in international organizations and other healthcare providers. Sensitization programs should be age-specific as well as occupation-based.

## 1.0 Introduction

Coronavirus disease 2019 (COVID-19) has posed unprecedented healthcare challenges globally.^1^ Without the control of the spread of the infections, COVID-19 would result in high morbidity and mortality, putting pressure on healthcare systems.^2^ The need to control the spread of COVID-19 infections is also associated with devastating financial effects and the likelihood of adverse long-term health consequences.^3^ The worldwide efforts towards stopping the spread of the pandemic focus on ensuring that majority of the global population acquire immunity, which could be through vaccination or infection.^2,4^ The other critical milestone is ensuring the widespread vaccination of the population to achieve herd immunity.^5^ The attainment of herd immunity, which is important in stopping the spread of COVID-19 infection negatively affected by vaccine hesitancy.^6^ In this study, vaccine hesitancy is defined as a deliberate delay or refusal to take any brand of the available and readily accessible vaccines.^4,5^

Healthcare workers play an important role in promoting vaccinations and in the prevention of the spread of infections.^5^ Due to the interaction with infected individuals, healthcare workers are at a higher risk of acquiring COVID-19 infections and could be the source of nosocomial infections.^7^ Therefore, healthcare workers play an important role in deterring the spread of infection or could be actively involved in increasing its spread. Evidence indicates that vaccine uptake among healthcare workers varies across different regions globally.^7^ Some researchers have reported less than 50 % willingness to get vaccinated among healthcare workers,^3,8^ while some evidence shows high willingness to get vaccinated.^1,2,4-6,9, 10^ There are various demographic factors that could influence acceptance of COVID-19 vaccines among healthcare workers. One of the factors is age where acceptance of COVID-19 vaccines is believed to be higher among older healthcare workers. ^2, 3, 7, 8, 10^ The gender of healthcare workers also could influence the acceptance of COVID-19 vaccines. However, there is contradicting evidence with other researchers^4,5^ indicating that being a male healthcare worker is significantly associated with the likelihood of accepting the vaccines. However, Kukreti and colleagues^3^ argue that gender is not a significant predictor of the willingness to receive vaccination among healthcare workers. Evidence also suggests that acceptance of COVID-19 vaccines among healthcare workers differs based on their hospital roles with acceptance being low among nurses and assistant nurses. ^4,6,7 11^ The other factors that could influence acceptance of COVID-19 vaccines among healthcare workers include level of education.^2^

The other issue that has been associated with COVID-19 vaccine hesitancy is trust.^12^ The fast-tracked manner in which the vaccines are produced also adds to increased vaccine hesitancy. Qattan and colleagues^5^ noted that 50.29% (N= 673) of the healthcare workers in Saudi Arabia were unwilling to get vaccinated unless the safety of the vaccine was confirmed. Gadoth and colleagues^10^ also reported vaccine hesitancy among 65.5% of the participants, which was due to concerns over safety.

It is evident from the reviewed evidence that there is variation in the level of acceptance of COVID-19 vaccines among the healthcare workers. It is also evident that demographic and social factors could influence the level of vaccine acceptance. Therefore, the aim of this study was to assess the influence of social trust and the demographic factors on the acceptance of COVID-19 vaccines among healthcare workers.

The twin island Republic of Trinidad and Tobago has an estimated population of 1.4 million^13^. The country reported its first case of SARS-CoV-2 on March 12, 2020^14^. Since then, public health measures such as border closures, social distancing, and mask-wearing have been implemented to limit the spread of the virus^15^. On February 17th, 2021, Trinidad and Tobago joined the global effort to control the pandemic through vaccination when the Ministry of Health embarked upon the Phase 1 rollout of its National COVID-19 Vaccination Program, with healthcare workers being among the first groups to receive the first doses of the vaccine, along with persons aged 60 years and over and persons with non-communicable diseases. By April 2021, subsequent phases (2 and 3) of the campaign offered frontline essential workers and the eligible public the opportunity to be inoculated. As of October 31st, 2021, approximately 43.0% (601,791) of the population were fully vaccinated (received all doses in a primary vaccine series)^16^.

Researchers in Trinidad and Tobago reported on the safety of the COVID-19 vaccine by examining the side-effects of the ChAdOx1 nCov-19 (Oxford, AstraZeneca COVID-19 vaccine) among healthcare workers^17^. The study demonstrated that the rate of occurrence of most local and systemic side-effects was less than 50%, corroborating the manufacturer’s claim that the vaccine is safe, with implications to reduce vaccine hesitancy through public health efforts. Other studies in Trinidad and Tobago have been limited to investigating COVID-19 patients’ epidemiological characteristics^18^ as well as laboratory predictors of COVID-19 admissions to ICU^19^. The most frequent comorbidities were found to be hypertension and diabetes mellitus, while the most prevalent symptoms were non-productive coughs and fevers^18^. As for laboratory factors, neutrophils, aspartate transaminase (AST), lactate dehydrogenase (LDH) and C-reactive protein (CRP) were suitable predictors of COVID-19 patients in need of ICU care(reference)^19^. Both studies allude to the unique characteristics of COVID-19 patients in Trinidad and Tobago and the greater need for research especially in this region.

## 2.0 Methods

### 2.1 Study setting and study design

This cross-sectional study involved the collection of data from 584 healthcare workers (HCWs) of the North Central Regional Health Authority (NCRHA), concerning their uptake, brand preferences, attitudes, and concerns, with respect to the COVID-19 vaccine. The NCRHA is one of four RHAs in Trinidad and comprises two of the eight counties in Trinidad and Tobago. At the time of this study, COVID-19 vaccination services were provided to the public at eleven (11) health centers, two (2) district health facilities, and four (4) mass vaccination sites in these counties. The NCRHA was selected as the setting for HCWs as it was the first RHA to distribute COVID-19 vaccines to HCWs at the outset of the country’s national vaccination program [20]. Data capture was conducted via the electronic distribution of a self-administered questionnaire to NCRHA HCWs. The survey remained open for responses from August 23rd, 2021 to October 31st, 2021. The anonymous responses were automatically collated via the online platform to which only the principal investigator had access. The collated responses were downloaded as a Microsoft Excel file by the principal investigator, and subsequently coded into an SPSS database and analyzed using IBM SPSS V.21 software.

The study protocol was reported following the Strengthening the Reporting of Observational Studies in Epidemiology (STROBE) guidelines for cross-sectional studies [21].

### 2.2 Study participants

A judgment sampling method was used to obtain the sample of HCWs for this study. The electronic questionnaire was distributed via email to all NCRHA HCWs of the following categories: medical practitioners, nursing personnel, veterinary surgeons, medical interns, dental interns, paramedics, and pharmacists. This list of healthcare workers was selected given that these categories of healthcare workers were granted power to administer COVID-19 vaccines during this period of public emergency by the President of Trinidad and Tobago and were therefore likely to be NCRHA healthcare workers directly involved in vaccine administration services at the RHA’s health centers, district health facilities or mass vaccination sites. Of the 1,351 NCRHA HCWs to whom the questionnaire was sent, 584 HCWs returned a completed questionnaire during the study period. Participation in this study was voluntary and HCWs received no form of financial remuneration in order to reduce the risk of response bias.

### 2.3. Instrument

The electronic self-administered questionnaire was prefaced with an informed consent form which explained that the survey was anonymous, that participation was voluntary, and explained the purpose of the study. The instrument (see supplementary file) consisted of four sections, modeled on relevant questions selected from validated questionnaires investigating the knowledge, attitudes, acceptance and concerns of HCWs and the wider population [22, 23, 24]. The first section gathered demographic data (age, gender, level of education, income level, profession). The second section asked about COVID-19 vaccine uptake, the third consisted of questions about COVID-19 vaccine brand preferences, while the fourth inquired about HCWs’ concerns with respect to the COVID-19 vaccine. The electronic questionnaire was pretested among twenty HCWs to obtain feedback on the clarity of questions and the length of time to complete the questionnaire. The questionnaire was finalized and subsequently distributed to the rest of the HCWs.

### 2.4. Outcome Measures

The main outcome measures of this study included the HCWs’ COVID-19 vaccine acceptance.

### 2.5. Ethics

This study was granted ethical approval by The North Central Regional Health Authority Ethics Committee, Trinidad, and The Ministry of Health of Trinidad and Tobago (3/13/441 Vol. II) Ethics Committee.

### 2.6. Analyses

#### Bivariate analysis

Bivariate analysis was carried out using Chi-square analysis of association. The bivariate analysis facilitated the determination of association between the respondents’ characteristics and the acceptance of the vaccine. Chi-square analysis of association was also used to determine the association between vaccine acceptance among healthcare workers and trust. The determination of the significant outcomes was carried out at p< 0.05.

#### Multinomial logistic regression

The independent association between the acceptance of the COVID-19 vaccines and healthcare workers’ characteristics and trust was established using multinomial logistic regression. The analysis approach was appropriate because the dependent variable had three categories. In this study, the category “ No, I am not willing to take any brand of the vaccine” was used as the reference category. The determination of the significant outcomes was carried out at p< 0.05. The analysis was carried out using SPSS.

## 3.0 Results

A total of 584 healthcare workers participated and 79.5% were female and 88.0% had received the COVID-19 vaccine. A high percentage (43.2 %) were aged between 25 and 34 years, while 36.8% were aged between 35 and 44 years. The participants were from different professionals with the majority (46.2%) being medical practitioners while 37.7% were registered nurses. The monthly income varied with 34.8% earning between 20,001 - 30,000 and 32.7% earning between 5,001 - 10,000. Most of the participants (64.7 %) had a bachelor’s degree and only 33.6% played a role in the vaccination process, which included patient screening, vaccine administration, patient observation, and collating of vaccine forms. The in-depth description of the sociodemographic attributes is shown in Table 1.

**Table 1:**
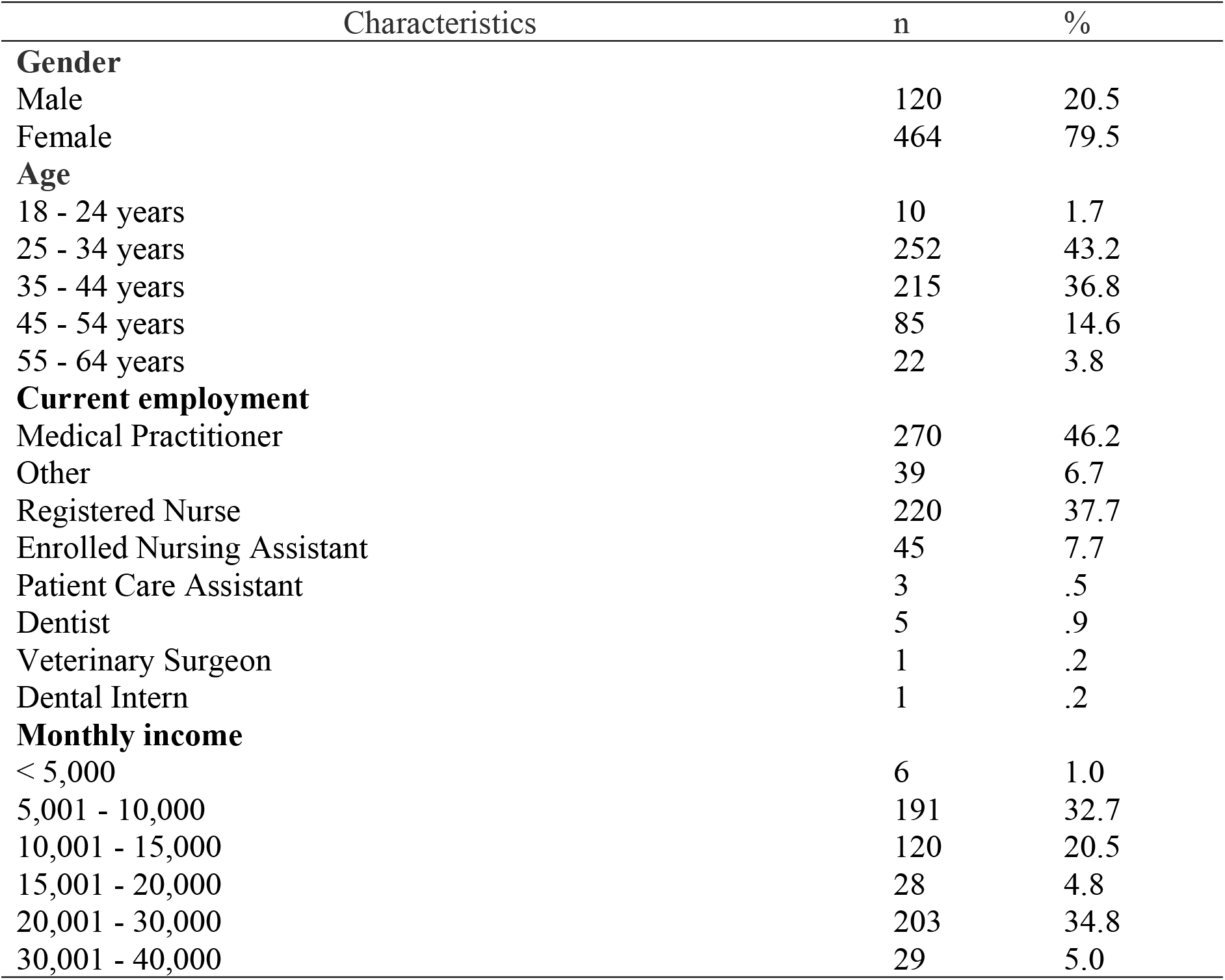
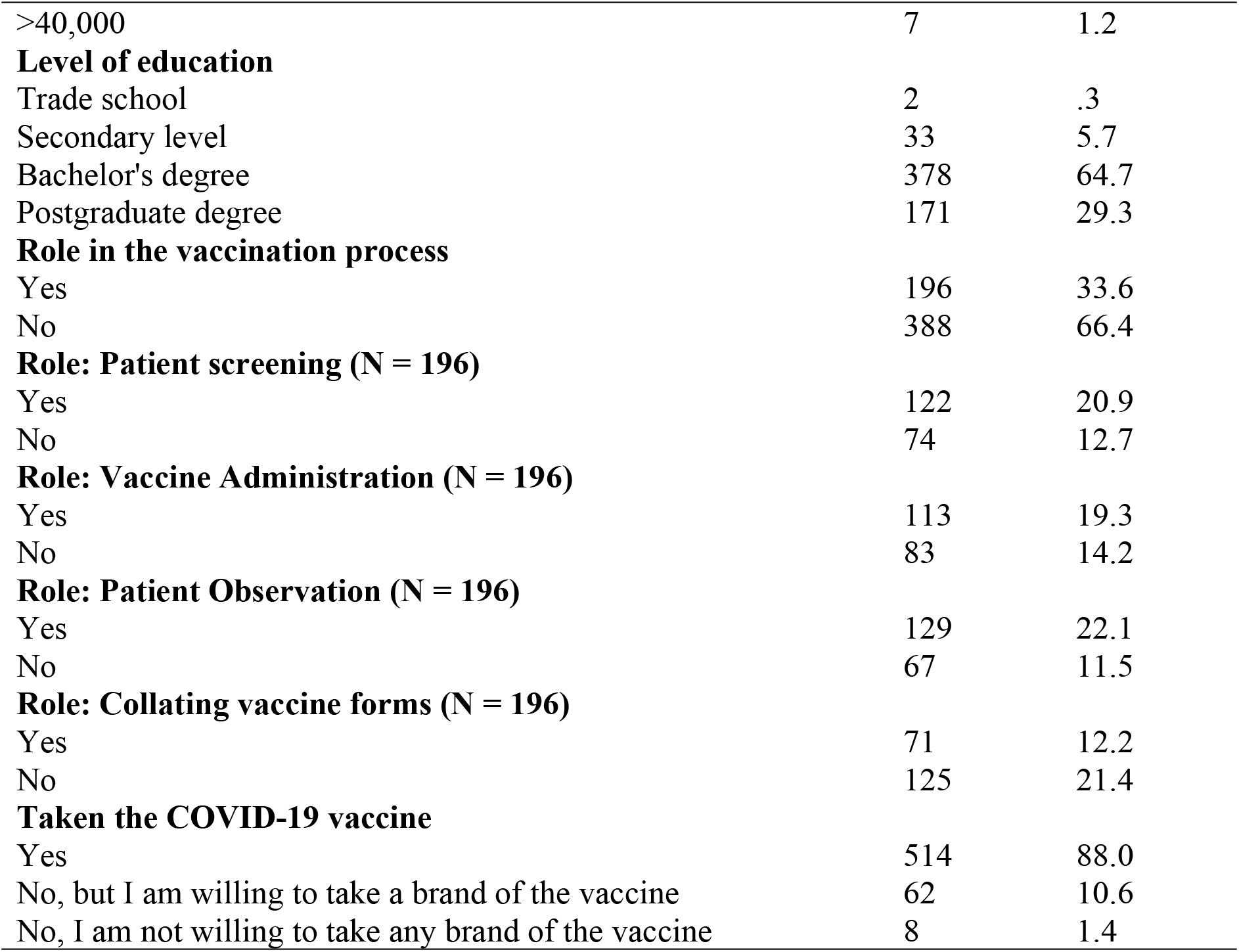
Demographic characteristics of the study participants (N = 584)

| Characteristics | n | % |
| --- | --- | --- |
| **Gender** |  |  |
| Male | 120 | 20.5 |
| Female | 464 | 79.5 |
| **Age** |  |  |
| 18 - 24 years | 10 | 1.7 |
| 25 - 34 years | 252 | 43.2 |
| 35 - 44 years | 215 | 36.8 |
| 45 - 54 years | 85 | 14.6 |
| 55 - 64 years | 22 | 3.8 |
| **Current employment** |  |  |
| Medical Practitioner | 270 | 46.2 |
| Other | 39 | 6.7 |
| Registered Nurse | 220 | 37.7 |
| Enrolled Nursing Assistant | 45 | 7.7 |
| Patient Care Assistant | 3 | .5 |
| Dentist | 5 | .9 |
| Veterinary Surgeon | 1 | .2 |
| Dental Intern | 1 | .2 |
| **Monthly income** |  |  |
| < 5,000 | 6 | 1.0 |
| 5,001 - 10,000 | 191 | 32.7 |
| 10,001 - 15,000 | 120 | 20.5 |
| 15,001 - 20,000 | 28 | 4.8 |
| 20,001 - 30,000 | 203 | 34.8 |
| 30,001 - 40,000 | 29 | 5.0 |
| >40,000 | 7 | 1.2 |
| **Level of education** |  |  |
| Trade school | 2 | .3 |
| Secondary level | 33 | 5.7 |
| Bachelor's degree | 378 | 64.7 |
| Postgraduate degree | 171 | 29.3 |
| **Role in the vaccination process** |  |  |
| Yes | 196 | 33.6 |
| No | 388 | 66.4 |
| **Role: Patient screening (N = 196)** |  |  |
| Yes | 122 | 20.9 |
| No | 74 | 12.7 |
| **Role: Vaccine Administration (N = 196)** |  |  |
| Yes | 113 | 19.3 |
| No | 83 | 14.2 |
| **Role: Patient Observation (N = 196)** |  |  |
| Yes | 129 | 22.1 |
| No | 67 | 11.5 |
| **Role: Collating vaccine forms (N = 196)** |  |  |
| Yes | 71 | 12.2 |
| No | 125 | 21.4 |
| **Taken the COVID-19 vaccine** |  |  |
| Yes | 514 | 88.0 |
| No, but I am willing to take a brand of the vaccine | 62 | 10.6 |
| No, I am not willing to take any brand of the vaccine | 8 | 1.4 |

Among the participants, 514 had taken the covid-19 vaccine (88.0%), 62 (10.6%) had not taken the vaccine but were willing to take a brand of the vaccine while eight (1.4%) had not taken the vaccine and were not willing to take any brand of the vaccine. Table 2 shows that the outcome of the Chi-square analysis of association between the respondents’ characteristics and the acceptance of the vaccine. A statistically significant association between the acceptance of vaccines and gender existed (p = 0.004) with a high number of females (87.5%) compared to males (12.5%) not having taken any vaccine and not willing to take any brand. However, it should be noted that the reported association between gender and acceptance of vaccines could be influenced by the disparity in the number of males included in the study. The acceptance of the vaccines was also associated with the participant’s age (p= 0.002) with the unwillingness to take the vaccines being reported among those aged 25 - 34 years (75.0%) and 35 - 44 years (25.0%). Vaccine acceptance was also associated with the current employment (p < 0.001), with the unwillingness to take the vaccines being reported among registered nurses (62.5%) and enrolled nurses (37.5%). A statistically significant association also existed between the monthly income and vaccine acceptance (p < 0.001) with the unwillingness to take the vaccine being reported only among those who earned between 10,001 - 15,000. Vaccine acceptance was also associated with the level of education (p < 0.001), with the unwillingness to take the vaccines being reported among those with Secondary level (25.0%) and Bachelor’s degree (75.0%). The participants’ role in the vaccination process was also statistically significantly associated with acceptance of vaccine (p = 0.003) with a high percentage of those unwilling to accept the vaccines being those who did not participate in the process (75%). A significant association existed between the acceptance of the vaccine and the participant’s role in patient screening (p = 0.011), vaccine administration (p = 0.018), patient observation (p = 0.005), and collating vaccine forms (p = 0.013) as shown in Table 2.

**Table 2:** Chi-square analysis of association between the respondents’ characteristics and the acceptance of the vaccine.

| Characteristic | Uptake of vaccines n (%) |  |  | P value |
| --- | --- | --- | --- | --- |
|  | Yes | No/I am willing | No/not willing |  |
| **Gender** |  |  |  |  |
| Male | 116 (22.6) | 3 (4.8) | 1 (12.5) | **0.004** |
| Female | 398 (77.4) | 59 (95.2) | 7 (87.5) |  |
| **Age** |  |  |  |  |
| 18 - 24 years | 5 (1.0) | 5 (8.1) | 0 | **0.002** |
| 25 - 34 years | 218 (42.4) | 28 (45.2) | 6 (75.0) |  |
| 35 - 44 years | 190 (37.0) | 23 (37.1) | 2 (25.0) |  |
| 45 - 54 years | 79 (15.4) | 6 (9.7) | 0 |  |
| 55 - 64 years | 22 (4.3) | 0 | 0 |  |
| **Current employment** |  |  |  |  |
| Medical Practitioner | 262 (51.0) | 8 (12.9) | 0 | < **0.001** |
| Other | 36 (7.0) | 3 (4.8) | 0 |  |
| Registered Nurse | 181 (35.2) | 34 (54.8) | 5 (62.5) |  |
| Enrolled Nursing Assistant | 26 (5.1) | 16 (25.8) | 3 (37.5) |  |
| Patient Care Assistant | 2 (0.4) | 1 (1.6) | 0 |  |
| Dentist | 5 (1.0) | 0 | 0 |  |
| Veterinary Surgeon | 1 (0.2) | 0 | 0 |  |
| Dental Intern | 1 (0.2) | 0 | 0 |  |
| **Monthly income** |  |  |  |  |
| < 5,000 | 4 (0.8) | 2 (3.2) | 0 |  |
| 5,001 - 10,000 | 144 (28.0) | 39 (62.9) | 8 (100) |  |
| 10,001 - 15,000 | 107 (20.8) | 13 (21.0) | 0 |  |
| 15,001 - 20,000 | 26 (5.1) | 2 (3.2) | 0 | < 0.001 |
| 20,001 - 30,000 | 198 (38.5) | 5 (8.1) | 0 |  |
| 30,001 - 40,000 | 28 (5.4) | 1 (1.6) | 0 |  |
| >40,000 | 7 (1.4) | 0 | 0 |  |
| **Level of education** |  |  |  |  |
| Trade school | 1 (0.2) | 1 (1.6) | 0 |  |
| Secondary level | 25 (4.9) | 6 (9.7) | 2 (25) | < 0.001 |
| Bachelor's degree | 319 (62.1) | 53 (85.5) | 6 (75.0) |  |
| Postgraduate degree | 169 (32.9) | 2 (3.2) | 0 |  |
| **Role in the vaccination process** |  |  |  |  |
| Yes | 185 (36.0) | 9 (14.5) | 2 (25.0) | 0.003 |
| No | 329 (64.0) | 53 (85.5) | 6 (75.0) |  |
| **Role: Patient screening** |  |  |  |  |
| Yes | 113 (22.0) | 7 (11.3) | 2 (25.0) | 0.011 |
| No | 72 (14.0) | 2 (3.2) | 0 |  |
| **Role: Vaccine Administration** |  |  |  |  |
| Yes | 106 (20.6) | 6 (9.7) | 1 (12.5) | 0.018 |
| No | 79 (15.4) | 3 (4.8) | 1 (12.5) |  |
| **Role: Patient Observation** |  |  |  |  |
| Yes | 124 (24.1) | 5 (8.1) | 0 | 0.005 |
| No | 61 (11.9) | 4 (6.5) | 2 (25.0) |  |
| **Role: Collating vaccine forms** |  |  |  |  |
| Yes | 67 (13.0) | 4 (6.5) | 0 | 0.013 |
| No | 118 (23.0) | 5 (8.1) | 2 (25.0) |  |

Table 3 shows the association between vaccine acceptance among healthcare workers and trust. The chi-square output indicates that there was a statistically significant association between the trust in the international television broadcast and the acceptance of vaccines (p = 0.014), with 62.5% of those unwilling to accept the vaccines being those having no trust or little trust in the international television broadcasting. Vaccine acceptance was also associated with the trust in the doctor/other healthcare professionals (p < 0.001), with 85.6% of those having taken the vaccines being those having moderate trust or a lot of trust in the doctor/other healthcare professionals. Table 3 also shows a statistically significant association between the trust in government agencies and the acceptance of vaccines (p < 0.001), with 87.5% of those unwilling to accept the vaccines being those having no trust or little trust in government agencies. It was also noted that the vaccine acceptance was also associated with the trust in the international organizations such as WHO and CDC (p < 0.001), with 91.0% of those having taken the vaccines being those having moderate trust or a lot of trust in the international organizations. As shown in Table 3, there was no statistically significant association between vaccine acceptance and the trust in national newspapers, international newspapers, national television broadcasts, national radio broadcasts, social media, and family and friends.

**Table 3:** Association between vaccine acceptance among healthcare workers and trust (N =584).

| Characteristic | Uptake of vaccines n (%) |  |  | P value |
| --- | --- | --- | --- | --- |
|  | Yes | No/I am willing | No/not willing |  |
| **Trust in: National newspapers** |  |  |  |  |
| No trust | 122 (23.7) | 17 (27.4) | 5 (62.5) | 0.105 |
| A little trust | 190 (37.0) | 23 (37.1) | 1 (12.5) |  |
| Moderate trust | 173 (33.7) | 22 (35.5) | 2 (25.0) |  |
| A lot of trust | 29 (5.6) | 0 | 0 |  |
| **Trust in: International newspapers** |  |  |  |  |
| No trust | 56 (10.9) | 11 (17.7) | 3 (37.5) | 0.116 |
| A little trust | 149 (29.0) | 22 (35.5) | 2 (25.0) |  |
| Moderate trust | 250 (48.6) | 23 (37.1) | 3 (37.5) |  |
| A lot of trust | 59 (11.5) | 6 (9.7) | 0 |  |
| **Trust in: National television broadcasts** |  |  |  |  |
| No trust | 77 (15.0) | 16 (25.8) | 2 (25.0) | 0.282 |
| A little trust | 168 (32.7) | 19 (30.6) | 4 (50.0) |  |
| Moderate trust | 221 (43.0) | 23 (37.1) | 2 (25.0) |  |
| A lot of trust | 48 (9.3) | 4 (6.5) | 0 |  |
| **Trust in: International television broadcasts** |  |  |  |  |
| No trust | 45 (8.8) | 10 (16.1) | 2 (25.0) | **0.014** |
| A little trust | 109 (21.2) | 21 (33.9) | 3 (37.5) |  |
| Moderate trust | 271 (52.7) | 25 (40.3) | 1 (12.5) |  |
| A lot of trust | 89 (17.3) | 6 (9.7) | 2 (25.0) |  |
| **Trust in: National radio broadcasts** |  |  |  |  |
| No trust | 115 (22.4) | 17 (27.4) | 4 (50.0) | 0.207 |
| A little trust | 175 (34.0) | 22 (35.5) | 4 (50.0) |  |
| Moderate trust | 188 (36.6) | 21 (33.9) | 0 |  |
| A lot of trust | 36 (7.0) | 2 (3.2) | 0 |  |
| **Trust in: Internet** |  |  |  |  |
| No trust | 46 (8.9) | 9 (14.5) | 1 (12.5) | 0.254 |
| A little trust | 130 (25.3) | 21 (33.9) | 3 (37.5) |  |
| Moderate trust | 254 (49.4) | 28 (45.2) | 3 (37.5) |  |
| A lot of trust | 84 (16.3) | 4 (6.5) | 1 (12.5) |  |
| **Trust in: Social media (Facebook, Instagram, WhatsApp, Twitter, TikTok & YouTube)** |  |  |  |  |
| No trust | 236 (45.9) | 25 (40.3) | 5 (62.5) | .173 |
| A little trust | 158 (30.7) | 25 (40.3) | 2 (25.0) |  |
| Moderate trust | 106 (20.6) | 12 (19.4) | 0 |  |
| A lot of trust | 14 (2.7) | 0 | 1 (12.5) |  |
| **Trust in: Your doctor/other healthcare professional** |  |  |  |  |
| No trust | 16 (3.1) | 9 (14.5) | 1 (12.5) | < 0.001 |
| A little trust | 58 (11.3) | 18 (29.0) | 3 (37.5) |  |
| Moderate trust | 246 (47.9) | 22 (35.5) | 3 (37.5) |  |
| A lot of trust | 194 (37.7) | 13 (21.0) | 1 (12.5) |  |
| **Trust in: Government Agencies** |  |  |  |  |
| No trust | 58 (11.3) | 17 (27.4) | 3 (37.5) | <0.001 |
| A little trust | 143 (27.8) | 24 (38.7) | 4 (50.0) |  |
| Moderate trust | 223 (43.4) | 16 (25.8) | 1 (12.5) |  |
| A lot of trust | 90 (17.5) | 5 (8.1) | 0 |  |
| **Trust in: International organizations (WHO, CDC, others)** |  |  |  |  |
| No trust | 12 (2.3) | 5 (8.1) | 1 (12.5) | < 0.001 |
| A little trust | 34 (6.6) | 11 (17.7) | 6 (75.0) |  |
| Moderate trust | 161 (31.3) | 28 (45.2) | 1 (12.5) |  |
| A lot of trust | 307 (59.7) | 18 (29.0) | 0 |  |
| **Trust in: Family and friends** |  |  |  |  |
| No trust | 144 (28.0) | 18 (29.0) | 4 (50.0) | 0.667 |
| A little trust | 202 (39.3) | 27 (43.5) | 1 (12.5) |  |
| Moderate trust | 134 (26.1) | 14 (22.6) | 2 (25.0) |  |
| A lot of trust | 34 (6.6) | 3 (4.8) | 1 (12.5) |  |

Multinomial logistic regression was carried out to determine the independent association between the acceptance of the COVID-19 vaccines and healthcare workers’ characteristics and trust. The model involved the use of the willingness to accept any brand of vaccine as the reference category. In Table 4, the outputs for the comparison categories, which were set to zero because of redundancy, were omitted. The predictors shown in Table 4 include the sex, profession, healthcare workers’ role during vaccination, trust, and healthcare workers’ age. Age, profession and trust are expressed as dichotomous variables. For variable trust, the new categories were No trust (included the responses no trust and a little trust) and trust (included moderate and a lot of trust). For the variable age, the categories included less than 35 years and 35 years and above. For the variable profession, the categories included nursing profession and healthcare workers of other professions. Table 4 shows that the odds of healthcare workers aged less than 35 years having taken the COVID-19 vaccine compared to those unvaccinated and unwilling to take any brand of the vaccine is significantly lower compared to the healthcare workers aged above 35 years (p = 0.02). Table 4 also indicates that the odds of having taken the COVID-19 vaccine compared to those unvaccinated and unwilling to take any brand of the vaccine is significantly higher among nurses compared to those of other profession (p = 0.01). The findings also indicate that the odds of having taken the COVID-19 vaccine compared to those unvaccinated and unwilling to take any brand of the vaccine were significantly lower among those who had no trust in international organizations (WHO, CDC, others) compared to those with trust (p = 0.001). The odds of being unvaccinated but willing to take any brand of the vaccine compared to those unvaccinated and unwilling to take any brand of the vaccine were significantly lower among those who had no trust in international organizations (WHO, CDC, others) compared to those with trust (p = 0.001). Table 4 also indicates that the odds of being unvaccinated but willing to take any brand of the vaccine compared to those unvaccinated and unwilling to take any brand of the vaccine were significantly lower among those who had no trust in doctor/other healthcare professional compared to those with trust (p = 0.04).

**Table 4:** Multinomial logistic regression indicating the independent association between the acceptance of the COVID-19 vaccines and healthcare workers’ characteristics and trust (N = 584).

|  |  | Yes, have taken the COVID-19 vaccine<sup>a</sup> |  | No, but I am willing to take any brand of the vaccine <sup>a</sup> |  |
| --- | --- | --- | --- | --- | --- |
|  |  | p-value | AOR (95% CI) | p-value | AOR (95% CI) |
| Age (Years) | Less than 35 | **.02** | .08 (.01-.68) | .06 | .13 (.01-1.07) |
|  | 35 and above | c | c | c | c |
| Sex | Male | .83 | .76 (.06-8.93) | .21 | .18 (.01-2.64) |
|  | Female | c | c | c | c |
| Profession | Nursing | **.01** | .09 (.01-.58) | .43 | .46 (.07-3.10) |
|  | Others | c | c | c | c |
| Role in the vaccination | Yes | .44 | 2.15 (.31-15.07) | .80 | .77 (.1-5.91) |
|  | No | c | c | . | . |
| Trust in: International television broadcasts | No trust | .85 | 1.23 (.15-10.03) | .58 | 1.83 (.21-15.88) |
|  | Trust | c | c | c | c |
| Trust in: Your doctor/other healthcare professional | No trust | .22 | 3.81 (.46-31.46) | **.04** | 10.66 (1.19-95.64) |
|  | Trust | c | c | c | c |
| Trust in: Government Agencies | No trust | .99 | .99 (.06-15.84) | .72 | 1.66 (.1-28.11) |
|  | Trust | c | c | c | c |
| Trust in: International organizations (WHO, CDC, others) | No trust | **.001** | .01 (.00-.10) | **.001** | .004 (.00-.11) |
|  | Trust | c | c | c | c |

Note. AOR = Adjusted Odds ratios CI =confidence interval. Cox & Snell $R^2 = 0.171$ ; Nagelkerke; $R^2 = 0.306$ ; McFadden $R^2 = 0.229$ ; <sup>a</sup> = compared to No, I am not willing to take any brand of the vaccine. c. This parameter is set to zero because it is redundant.

## 4.0 Discussion

Based on the outcome of this study, a low percentage of healthcare workers show vaccine hesitancy. The outcome contradicts studies that have shown vaccine acceptance among healthcare workers to be less than 50 %.^3,8^ Kukreti and colleagues^3^ assessed the acceptance of vaccines among 500 health care workers in Taiwan. The researchers observed that only 23.4% of the healthcare workers showed willingness to be vaccinated. The outcome corroborates the observation made by Gagneux-Brunon and colleagues^4^ and Kuter and colleagues^2^ which showed high percentage of vaccine acceptance. The study also supports the findings by Gadoth and colleagues^10^ that showed that among 540 American healthcare workers, 46.9% perceived COVID-19 vaccines to offer protection against infections. Gagneux-Brunon and colleagues^4^ also reported that among 2047 French health care workers, COVID-19 vaccine acceptance was 75%. Maltezou and colleagues^6^ reported the acceptance of COVID-19 vaccines among 51.1% of the 1571 healthcare workers from Greece. Qattan and colleagues^5^ noted that among 673 healthcare workers in Saudi Arabia, 50.52% showed a willingness to get vaccinated. Kuter and colleagues^2^ reported that 63.7% of 12034 healthcare workers selected from Philadelphia, United States showed readiness to accept COVID-19 vaccines. The reported high vaccine acceptance also supports the findings by Meyer and colleagues^8^ also noted that among 16292 American healthcare workers, only 55.3% showed a willingness to receive COVID-19 vaccines.

Although the outcome of the study indicated that a high percentage of females compared to males showed vaccine hesitancy, the logistic regression outcome showed that gender does not predict the uptake of COVID-19 vaccines among healthcare workers. The outcome of the study contradicts the observations made by other researchers. ^4-6^ Qattan and colleagues^5^ reported that gender significantly predicts the uptake of vaccines by healthcare workers with males having higher acceptance. Wang and colleagues^8^ also observed that being a male healthcare worker significantly predicted the intention to accept the vaccines. Kuter and colleagues^2^ and Maltezou and colleagues^6^ also reported that being a male healthcare worker was significantly associated with the likelihood of accepting the vaccines. Gagneux-Brunon and colleagues^4^ also reported that male healthcare workers were inclined towards COVID-19 vaccine acceptance. However, the reported findings corroborate the observation made by Kukreti and colleagues^3^ which also showed that gender does not significantly predict the willingness to receive COVID-19 vaccine among healthcare workers. The disparity between the outcome of this study and those of previous researchers could be associated with an unequal sample size between males and females since males only made up 20.5 % of the study sample.The outcome of the study supports the view that the age of the healthcare workers is associated with the acceptance of COVID-19, which contradicts observations made by Kukreti and colleagues^3^. Other researchers have also shown age to have no significant influence on the prediction of willingness to get vaccinated among healthcare workers.^5,8^ The outcome of the study further indicated the age of the healthcare workers predict vaccine acceptance. The observation corroborates the conclusions made by Raftopoulos and colleagues^7^ following the assessment of vaccine acceptance among 223^8^ health care workers from Greece and the Republic of Cyprus. Raftopoulos and colleagues^7^ noted that age significantly predicts the willingness of healthcare workers to be vaccinated. The reported findings also support the observations made by Wang and colleagues^1^ who noted that individuals who were younger than 50 years were more willing to get vaccinated. Gadoth and colleagues^10^ also reported that older healthcare workers (50 years or older) were significantly more likely to accept vaccines. Similar observations were also made by Kuter and colleagues^2^.Various researchers have also shown that younger age predicts vaccine hesitancy.^1,7^

The findings of the study also indicate that the healthcare profession is associated with COVID-19 vaccines among healthcare workers. According to this study being a nurse is significantly associated with vaccine uptake compared to other professions. The findings corroborate the observations made by Shaw and colleagues^11^ regarding the difference in the acceptance of COVID-19 vaccines among healthcare workers based on their hospital roles. However, the findings also support the conclusions made by Gagneux-Brunon and colleagues^4^ showing that nurses and assistant nurses were less likely to accept COVID-19 vaccination.

The outcome of the study regarding the influence of social trust indicated that the trust in care provider and the international organizations such as WHO and CDC are associated with vaccine acceptance. Rozek and colleagues ^25^ also noted the trust in healthcare providers and scientists is vital in enhancing the acceptance of vaccines. According to this study, the trust in government agencies does not significantly predict uptake of vaccines among healthcare workers, which contradicts previous researchers. ^25, 26^ Park and colleagues^26^ also reported that the low trust in government agencies is associated with a low level of COVID-19 vaccine acceptance.

### 4.1 Study limitations

One of the limitations in study is the unequal sample size. The number of male participants was smaller compared to that of females, which made the determination of the influence of gender problematic. Given on the reported level of vaccine hesitancy of 1.4%, it is likelihood that the chosen population was not appropriate for evaluating the factors associated with hesitancy.

## 5.0 Conclusion

A low percentage of healthcare workers showed vaccine hesitancy. The study indicates that age, profession, and trust in international organizations and other healthcare providers are associated with the uptake of COVID-19 vaccines among healthcare workers. However, gender of the healthcare workers is not associated vaccine acceptance. Based on the outcome of the study, efforts towards enhanced vaccine acceptance among healthcare workers should take into consideration the age, profession, and the trust in international organizations and other healthcare providers.

## Data Availability

The data that support the findings of this study are available from the corresponding author, C.G., upon reasonable request.

## 6.0 Declarations

### 6.2 Authors’ Contributions

CDG and DV were responsible for data analysis, with intellectual contributions from DT. CDG and DV drafted the article. All authors contributed to the conception and design of the paper, interpretation of data, and critical revisions contributing to the intellectual content and approval of the final version of the manuscript.

### 6.4 Ethics Approvals

The North Central Regional Health Authority Ethics Committee – approval granted The Ministry of Health of Trinidad and Tobago Ethics Committee – approval granted

